# Gut microbiome alterations in fecal samples of treatment-naïve *de novo* Parkinson’s disease patients

**DOI:** 10.1101/2022.02.18.22270887

**Authors:** Jeffrey M Boertien, Kirsi Murtomäki, Pedro AB Pereira, Sygrid van der Zee, Tuomas H Mertsalmi, Reeta Levo, Tanja Nojonen, Elina Mäkinen, Elina Jaakkola, Pia Laine, Lars Paulin, Eero Pekkonen, Valtteri Kaasinen, Petri Auvinen, Filip Scheperjans, Teus van Laar

## Abstract

Gut microbiota alterations in Parkinson’s disease (PD) have been found in several studies and are suggested to contribute to the pathogenesis of PD. However, previous results could not be adequately adjusted for a potential confounding effect of PD medication and disease duration, as almost all PD participants were already using dopaminergic medication and were included several years after diagnosis. Here, the gut microbiome composition of treatment-naïve *de novo* PD subjects was assessed compared to healthy controls (HC) in two large independent case-control cohorts (n=136 and 56 PD, n=85 and 87 HC), using 16S-sequencing of fecal samples. Relevant variables such as technical batches, diet and constipation were assessed for their potential effects.

Overall gut microbiome composition differed between PD and HC in both cohorts, suggesting gut microbiome alterations are already present in *de novo* PD subjects at the time of diagnosis, without the possible confounding effect of dopaminergic medication. Although no differentially abundant taxon could be replicated in both cohorts, multiple short chain fatty acids (SCFA) producing taxa were decreased in PD in both cohorts. In particular, several taxa belonging to the family Lachnospiraceae were decreased in abundance. Fewer taxonomic differences were found compared to previous studies, indicating smaller effect sizes in *de novo* PD.

**Trial registration:** DUPARC: NCT04180865

NMDAT: NCT02650843

## Introduction

Parkinson’s disease (PD) is a neurodegenerative disorder, clinically characterized by motor symptoms like bradykinesia, rigidity and tremor.^1^ Non-motor symptoms can precede the cardinal motor symptoms by years.^2^ In particular, constipation is one of the earliest non-motor manifestations of PD and can occur up to twenty years before diagnosis.^3^ The occurrence of non-motor symptoms can be used to identify probable prodromal PD subjects and predict conversion to motor PD.^4^ Complementary to the early gastrointestinal symptomatology, alpha-synuclein (aSyn) pathology, gut inflammation and increased gut permeability are present in the early and prodromal stages of the disease.^5,6^ In addition, ascending denervation along the vagal nerve is suggested in a subgroup of PD subjects, suggesting a gastrointestinal origin in a subtype of PD.^7^

The relation between gut health and PD has led to an increased interest in the putative role of the gut microbiome in PD. Several preclinical studies found evidence that gut microbiota may impact PD pathology via for instance cross-seeding of aSyn or inflammatory signaling through microbial metabolites, such as short chain fatty acids (SCFA).^8–10^ In addition, over fifteen case-control studies have found alterations in gut microbiome composition in PD.^11–15^ Though some associations could be robustly replicated, various results were inconsistent across several studies, in part due to differences in study population and methodology.

Of particular concern is the fact that almost all participants of previous studies were included several years after diagnosis and were already using dopaminergic medication.^11^ Therefore, a possible PD related effect cannot adequately be distinguished from a potential confounding effect of the dopaminergic medication. Levels of Lachnospiraceae, *Bifidobacterium* and *Lactobacillus* have indeed been correlated to levodopa dosage.^14,16,17^ Additionally, a recent mendelian randomization study suggested the association between PD and *Bifidobacterium* to be based on reverse causation.^18^ Also Catechol-O-methyltransferase (COMT) inhibitors have been associated with gut microbiome changes.^17^ Moreover, possible microbial biomarkers cannot be assessed for their potential as a diagnostic marker, as data at the time of diagnosis is lacking, and longer disease duration is associated with larger differences in gut microbiome composition in PD.^19^

In addition to established PD cases, gut microbiome changes have been described in probable prodromal PD subjects, with two studies reporting changes comparable to PD, and a third study linking gut microbiome changes to specific prodromal symptoms rather than overall prodromal risk.^20–22^ Polysomnography proven REM sleep behavior disorder (RBD) is the prodromal symptom with the highest predictive value for conversion to PD.^23^ The recently proposed dichotomy of body-first and brain-first PD suggests a possible gastrointestinal (body-first) origin of PD in subjects with RBD at the time of diagnosis and during the prodromal stages of the disease.^7^ Moreover, RBD is proposed to co-occur with other prodromal autonomic symptoms, such as constipation, in the body-first subtype.^24^ Therefore, results found in probable prodromal subjects might only reflect a particular subtype of PD.

There is still a large unmet need to assess the gut microbiome of treatment-naïve *de novo* PD subjects at the time of diagnosis, without the confounding effect of dopaminergic medication. Currently, the largest gut microbiome assessment of a treatment-naïve *de novo* PD cohort consists of a subcohort of 39 subjects by Barichella et al.^19^ Compared to already treated, more advanced PD subjects, the gut microbiome composition was less different from that of HC and only showed the family Lachnospiraceae and two of its genera (*Roseburia* and an unclassified genus) to be differentially abundant compared to HC.

The current gut microbiome study concerns two large, independent, treatment-naïve cohorts from the Netherlands (NL cohort) and Finland (FIN cohort). The gut microbiome of fecal samples was investigated using 16S rRNA gene sequencing. Both cohorts comprise both PD and HC subjects from the same geographical area. Both cohorts were analyzed separately due to their different geographical origin and the use of different methodologies for stool sample collection and DNA extraction. Relevant variables, such as technical batches, diet and constipation, were assessed for their potential effects on the relationship between PD status and gut microbiome composition. Subsequently, a thorough first investigation of the gut microbiome of two large treatment-naïve *de novo* PD cohorts could be conducted.

## Results

### Clinical characteristics

For the NL cohort, 136 PD subjects and 85 HC could be included. The PD and HC groups were similar in terms of age and BMI, but differed in terms of the sex ratio (p=0.026). In the FIN cohort 56 PD subjects and 87 HC were included. There was no statistically significant difference in age, sex and BMI between the FIN cohort PD and HC groups. As expected, gastrointestinal dysfunction was higher in PD compared to HC, with lower Bristol Stool Chart scores for stool consistency and lower stool frequency, indicating harder stool consistency and increased levels of constipation. However, these associations were statistically significant only in the NL cohort. An overview of the clinical characteristics is provided in table 1.

**Table 1.** Clinical characteristics

|  | NL cohort |  |  | FIN cohort |  |  |
| --- | --- | --- | --- | --- | --- | --- |
|  | PD | HC | p-value | PD | HC | p-value |
| n | 136 | 85 | - | 56 | 87 | - |
| Age, mean ±sd | 66.0 ±8.86 | 65.9 ±9.69 | 0.91 | 64.9 ±10.77 | 67.1 ±7.3 | 0.18 |
| Female, n(%) | 41 (30%) | 39 (46%) | 0.026 | 27 (48%) | 43 (49%) | 1.0 |
| BMI, mean ±sd | 26.5 ±4.46 | 26.1 ±3.73 | 0.52 | 26.3 ±5.62 | 26.6 ±3.98 | 0.76 |
| Stool consistency, mean ±sd | 3.40 ±0.94 | 3.85 ±0.85 | 3.5E-04 | 3.21 ±0.87 | 3.42 ±0.81 | 0.15 |
| Stool frequency, median [IQR] | 1 [0.82-1.29] | 1.17 [1-1.73] | 2.0E-05 | 1 [0.86-1.57] | 1.14 [1-1.57] | 0.13 |
| Gastric anacidic medication, n(%) | 32 (24%) | 14 (16%) | 8.0E-03 | 8 (14%) | 3 (3%) | 0.025 |
| Constipation medication, n(%) | 16 (12%) | 1 (1%) | 2.7E-04 | 6 (11%) | 7 (8%) | 0.77 |
| MDS-UPDRSIII total, mean ±sd | 31.8 ±11.63 | - | - | 33.8 ±13.03 | - | - |
| Hoehn and Yahr, median [IQR] | 2 [1-2] | - | - | 2 [1-2] | - | - |
| Motor symptom duration in months, mean ±sd | 21.6 ±14.36 | - | - | 20.9 ±14.29 | - | - |

NL cohort, Dutch case-control cohort; FIN cohort, Finnish case-control cohort; PD, Parkinson's disease; HC, healthy control; p-value, nominal p-value; n, number of participants; sd, standard deviation; BMI, body mass index; stool consistency, average stool consistency score according to the Bristol Stool Chart; stool frequency, average stool frequency per day; IQR, interquartile range; MDS-UPDRS, Movement Disorders Society Unified Parkinson's Disease Rating Scale; motor symptom duration in months estimated retrospectively by participants counting.

### Variable selection

Given the primary aim to describe gut microbiome changes for the first time in treatment-naïve de novo PD subjects, all clinical and technical metadata were investigated for their potential to influence the effect of PD status on gut microbiome composition. The variables age, sex, BMI, stool consistency and DNA extraction batch were selected a priori as potentially relevant nuisance variables. To ensure a large effect of another variable on the relation between PD status and gut microbiome composition would not be missed, all nuisance variables were screened for their potential effect using a PERMANOVA in a model with PD status:

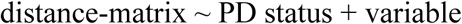

Variables that shifted the explained variance of PD status (R2) by at least ten percent compared to the univariable explained variance of PD status (distance-matrix ∼ PD status), were added to the list of relevant model covariates (Supplementary Table 1). Subsequently, the selection of relevant covariates was investigated for collinearity with PD status. Variables with a generalized variance inflation factor (GVIF) ≥2 with PD status were excluded. In the overall model, a GVIF of 3 was used as threshold for collinearity, excluding variables with a higher GVIF or one of several variables that only showed collinearity amongst each other (Supplementary Table 2). This resulted in the following models for the multivariate analysis of overall gut microbiome composition.

For the NL cohort: distance-matrix ∼ PD status + age + sex + BMI + stool consistency + NMSQ constipation + stool frequency + DNA extraction batch

For the FIN cohort: distance-matrix ∼ PD status + age + sex + BMI + stool consistency + CSI total score + strained defecation + number of sequences

For the differential abundance analysis, covariates were selected that showed a significant relationship with the overall gut microbiome composition in the final model with non-stringent p < 0.1, resulting in the following two models.

For the NL cohort: taxon ∼ PD status + BMI + stool consistency + stool frequency For the FIN cohort: taxon ∼ PD status + stool consistency + number of sequences

### Overall microbiome composition

Within-sample diversity (i.e. alpha diversity) indices indicated opposite effects in the two cohorts (Figure 1A). In the NL cohort, alpha diversity was higher in PD subjects with statistically significant differences in observed richness and Chao1 (nominal p-values < 0.05, uncorrected Mann-Whitney U test) and a statistically non-significant increase in the Shannon alpha diversity index. In contrast, alpha diversity was lower in the FIN cohort, with statistically significant differences in observed richness, Chao1, Shannon and inverse Simpson (nominal p-values < 0.01, uncorrected Mann-Whitney U test).

**Figure 1.**
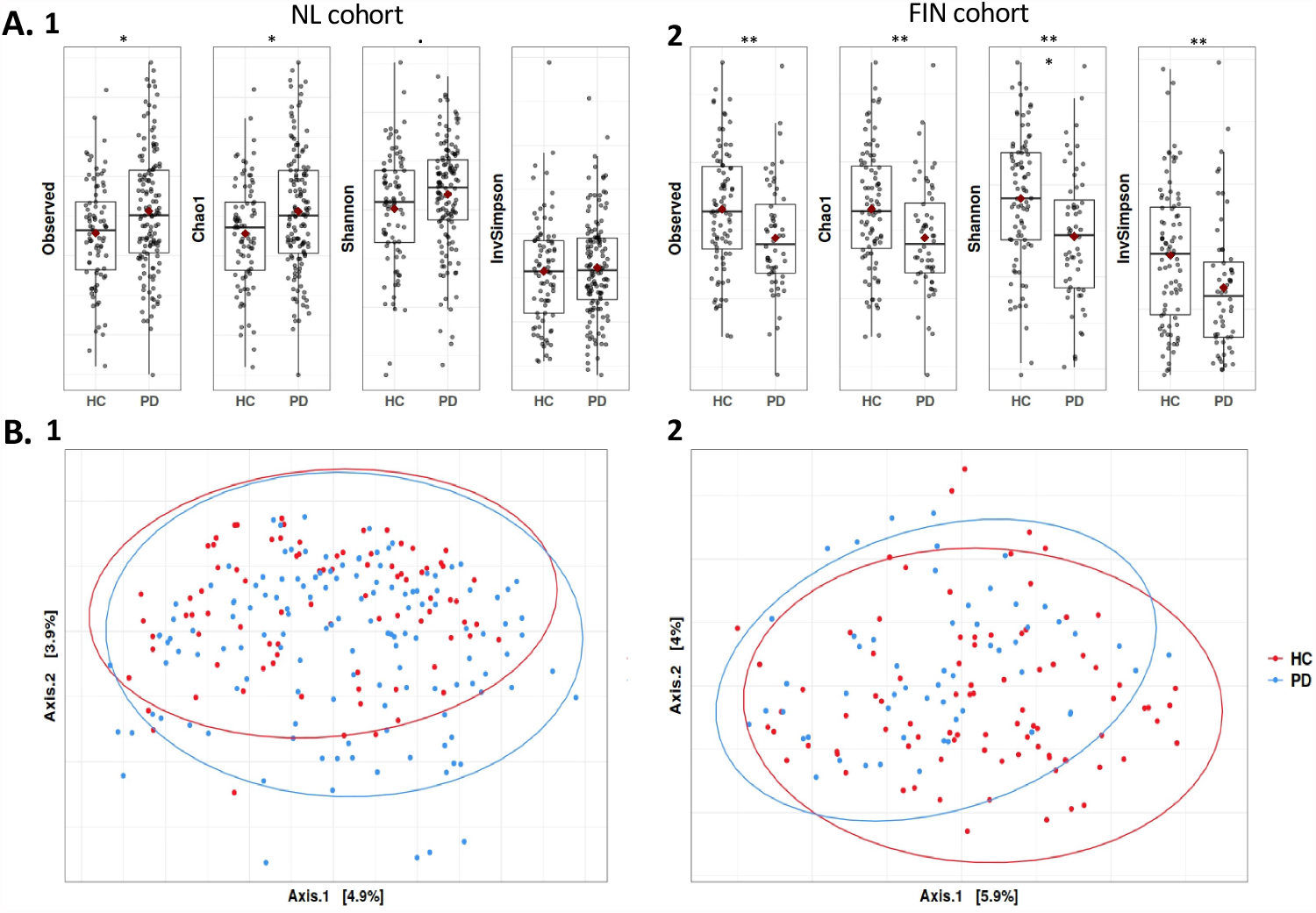
Intra-sample (alpha diversity) and inter-sample differences in microbial community structure between PD and HC with each point representing one sample. A) Alpha diversity indices indicate increased intra-sample diversity in PD in 1) the NL cohort, whereas intra-sample diversity in PD is reduced in 2) the FIN cohort. Univariable, uncorrected (Mann-Whitney U test) p-values are 0.016, 0.016, 0.060 and 0.56 for the NL cohort and 4.3E-03, 4.2E-03, 7.8E-04 and 3.9E-03 for the FIN cohort for, respectively, observed richness, Chao1, Shannon and Inverse Simpson. B) Inter-sample differences in microbial community structure were visualized using a principal component analysis of the Aitchinson distance, indicating a statistically significant differences between PD and HC with p=6.4E-03 and p=4.0E-04 for 1) the NL cohort and 2) the FIN cohort, respectively. Sample sizes: 136 PD and 85 HC for the NL cohort and 56 PD and 87 HC in the FIN cohort.. p < 0.1; * p < 0.05; ** p < 0.01; *** p < 0.001; NL cohort, Dutch case-control cohort; FIN cohort, Finnish case-control cohort; PD, Parkinson’s disease; HC, healthy control.

Overall gut microbiome composition (i.e. beta diversity) showed a large overlap between PD and HC (Figure 1B). PERMANOVA revealed univariable statistically significant differences between PD and HC in both cohorts: p=6.4E-03 and p=4.0E-04 for the NL and FIN cohort respectively (Table 2). After adjusting for selected covariates (see variable selection) a statistically significant difference between PD and HC remained, with p=0.030 and p=0.016 for the NL and FIN cohort respectively. When assessing differences in overall gut microbiome, using (non-compositional) Bray-Curtis distances, the results were similar, although no statistically significant difference between PD and HC was found in the NL cohort when adjusting for covariates (p=0.17).

**Table 2.** Comparisons of overall gut microbiome composition between PD and HC

|  |  | NL cohort |  |  |  | FIN cohort |  |  |  |
| --- | --- | --- | --- | --- | --- | --- | --- | --- | --- |
|  |  | Aitchinson |  | Bray-Curtis |  | Aitchinson |  | Bray-Curtis |  |
|  |  | var % | p-value | var % | p-value | var % | p-value | var % | p-value |
| **Univariable** |  |  |  |  |  |  |  |  |  |
|  | **group** | 0.62 | 6.4E-03 | 0.74 | 9.1E-03 | 1.18 | 4.0E-04 | 1.29 | 1.1E-03 |
| **Adjusted for confounders** |  |  |  |  |  |  |  |  |  |
|  | **group** | 0.57 | 0.030 | 0.54 | 0.17 | 0.92 | 0.016 | 1.03 | 0.020 |
|  | **age** | 0.70 | 4.0E-04 | 0.90 | 3.0E-04 | 0.63 | 0.85 | 0.64 | 0.66 |
|  | **sex** | 0.49 | 0.25 | 0.61 | 0.067 | 0.77 | 0.17 | 0.68 | 0.55 |
|  | **BMI** | 0.81 | <1.0E-04 | 0.99 | 3.0E-04 | 0.77 | 0.18 | 0.81 | 0.21 |
|  | **stoolconsistency** | 0.56 | 0.033 | 0.62 | 0.050 | 0.83 | 0.073 | 0.82 | 0.19 |
|  | **NMSQ constipation** | 0.52 | 0.10 | 0.55 | 0.15 | - | - | - | - |
|  | **stoolfrequency** | 0.57 | 0.027 | 0.62 | 0.056 | - | - | - | - |
|  | **DNA extraction batch** | 6.50 | 0.37 | 6.72 | 0.26 | - | - | - | - |
|  | **CSI total score** | - | - | - | - | 0.74 | 0.27 | 0.74 | 0.36 |
|  | **Strained defecation** | - | - | - | - | 0.69 | 0.52 | 0.66 | 0.61 |
|  | **Number of sequences** | - | - | - | - | 0.87 | 0.034 | 0.77 | 0.30 |

Differences in overall gut microbiome composition between PD and HC were assessed in a univariable analysis, and in a multivariable analysis with adjustment for selected variables. Each variable was adjusted for all other variables in the multivariable analysis (marginal testing). In addition to Aitchinson distances, the analyses were also performed with Bray-Curtis distances. Sample sizes for the univariable analyses were 136 PD and 85 HC for the NL cohort and 56 PD and 87 HC in the FIN cohort. Sample sizes for the multivariable analyses were 131 PD and 84 HC for the NL cohort and 54 PD and 87 HC for the FIN cohort. For the analyses based on Bray-Curtis distances, four additional PD and one additional HC sample were excluded from the NL cohort and one additional PD sample was excluded from the FIN cohort, due to insufficient number of sequences after rarefaction. P-values were based on 10,000 permutations, making the lowest possible p-value 9.999E-05, written as <1.0E-04 in above table. NL cohort, Dutch case-control cohort; FIN cohort, Finnish case-control cohort; PD, Parkinson's disease; HC, healthy control; BMI, body mass index; NMSQ, Non-Motor Symptom Questionnaire; CSI, Constipation Severity Instrument; strained defecation, level of straining during defecation measured in FIN cohort stool diary; number of sequences, number of sequences per sample.

### Differential abundance

Differential abundance analyses revealed several statistically significant differentially abundant taxa at the ASV, genus and family level (Table 3). Only one taxon at the ASV level, belonging to the Lachnospiraceae *GCA-900066575* genus, was identified as differentially abundant by both ANCOM and DESeq2 in the FIN cohort. None of the differentially abundant taxa were shared between the NL and FIN cohort. However, some similarity between the two cohorts can be discerned. Both at the ASV and genus level several taxa belonging to the Lachnospiraceae family were reduced in abundance in PD, in particular strains belonging to the Lachnospiraceae genus *Roseburia*. At the family level, Lachnospiraceae was reduced in PD in both cohorts, although this difference was not statistically significant after correction for multiple testing in DESeq2 nor identified as differentially abundant in ANCOM (Supplementary Table 5). Relative abundances of genera and families that were identified as statistically significant differentially abundant in one of the two cohorts, are depicted in Figure 2.

**Table 3.** Differentially abundant taxa between PD and HC

| Level | Family | Genus | Species | NL cohort |  |  |  |  | FIN cohort |  |  |  |  |
| --- | --- | --- | --- | --- | --- | --- | --- | --- | --- | --- | --- | --- | --- |
|  |  |  |  | baseMean | log2 Fold Change | DESeq2 (FDR) | ANCOM (W) | ANCOM significant | baseMean | log2 Fold Change | DESeq2 (FDR) | ANCOM (W) | ANCOM significant |
| ASV | Lachnospiraceae | Lachnospiraceae NK4A136 group | - | 74.61 | -1.61 | 1.0 | 285 | yes | 102.99 | 0.20 | 1.0 | 0 | no |
| ASV | Lachnospiraceae | Lachnoclostridium | - | 90.76 | -0.82 | 1.0 | 313 | yes | 99.74 | -1.24 | 1.0 | 216 | no |
| ASV | Lachnospiraceae | Roseburia | - | 100.23 | -1.46 | 1.0 | 274 | yes | 15.64 | -2.92 | 0.45 | 207 | no |
| ASV | Lachnospiraceae | Roseburia | inulinivorans | 266.76 | -0.14 | 1.0 | 153 | no | 207.61 | -1.57 | 1.0 | 258 | yes |
| ASV | Lachnospiraceae | Lachnoclostridium | edouardi | 83.60 | -0.48 | 1.0 | 0 | no | 83.08 | -1.44 | 1.0 | 252 | yes |
| ASV | Lachnospiraceae | Lachnospiraceae UCG-004 | - | 39.01 | -0.33 | 1.0 | 0 | no | 65.47 | -1.84 | 0.67 | 252 | yes |
| ASV | Lachnospiraceae | GCA-900066575 | - | 17.16 | -0.23 | 1.0 | 0 | no | 14.32 | -3.50 | **4.4E-03** | 274 | yes |
| ASV | Oscillospiraceae | Colidextribacter | - | 10.45 | 0.24 | 1.0 | 0 | no | 16.60 | -1.27 | 1.0 | 248 | yes |
| ASV | Christensenellaceae | Christensenellaceae R-7 group | - | 8.41 | 0.59 | 1.0 | 6 | no | 23.85 | 1.58 | 1.0 | 279 | yes |
| Genus | Lachnospiraceae | Lachnospiraceae NK4A136 group | - | 367.11 | -0.43 | 0.99 | 88 | yes | 407.63 | -0.33 | 1.0 | 0 | no |
| Genus | Rikenellaceae | Rikenellaceae RC9 gut group | - | 157.67 | 1.67 | 0.99 | 89 | yes | 136.94 | -0.25 | 1.0 | 0 | no |
| Genus | Lachnospiraceae | Roseburia | - | 896.86 | -0.14 | 0.99 | 73 | no | 588.97 | -0.50 | 1.0 | 82 | yes |
| Genus | Butyricicoccaceae | Butyricicoccus | - | 187.67 | -0.21 | 0.99 | 66 | no | 147.39 | -0.95 | **0.033** | 49 | no |
| Family | Veillonellaceae | - | - | 842.30 | -1.62 | 0.99 | 42 | yes | 715.05 | -0.29 | 0.94 | 0 | no |
| Family | Butyricicoccaceae | - | - | 221.57 | -0.26 | 0.99 | 18 | no | 164.48 | -0.98 | **2.6E-03** | 22 | no |
| Family | Eggerthellaceae | - | - | 120.86 | -0.06 | 0.99 | 0 | no | 38.92 | 1.08 | **0.022** | 9 | no |

Differentially abundant taxa between PD and HC detected using DESeq2 and/or ANCOM in at least one of the two cohorts. Both analyses were adjusted for variables with $p < 0.1$ in the comparison of overall gut microbiome compositions (age, BMI, stool consistency and stool frequency in the NL cohort; stool consistency and number of reads in the FIN cohort). Only samples for which complete metadata concerning the selected confounders was available, were included in the differential abundance analyses, resulting in 131 PD and 84 HC subjects for the NL cohort and 55 PD and 87 HC subjects for the FIN cohort.
NL cohort, Dutch case-control cohort; FIN cohort, Finnish case-control cohort; PD, Parkinson's disease; HC, healthy control; BMI, body mass index; baseMean, mean of all normalized counts provided by DESeq2; log2 Fol Change, a negative change indicates lower abundance in PD compared to HC; DESeq2, Differential Expression analysis for Sequence count data; FDR, False Discovery Rate corrected p-value; ANCOM, ANalysis of Composition of Microbiomes; W, W statistic indicating the number of taxa relative to which the presented taxon was differentially abundant (maximum values for W were 371, 124 and 45 for the NL cohort and 354, 113 and 46 for the FIN cohort at the ASV, Genus and Family level, respectively); ANCOM significant, indicates whether the taxon is detected as significantly differentially abundant using a detection threshold of 0.7; ASV, Amplicon Sequence Variant.

**Figure 2.**
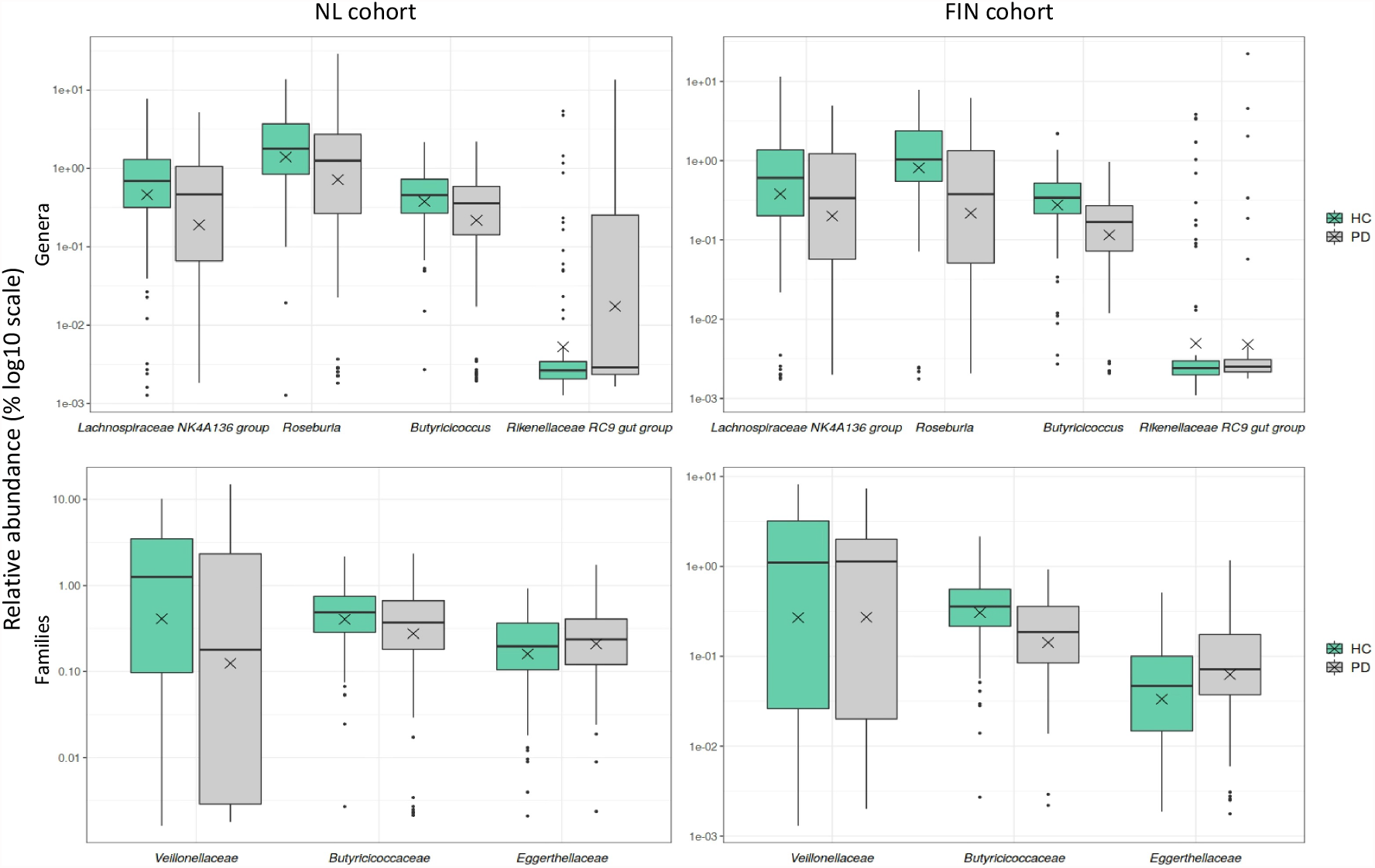
Relative abundances of genera and families identified as differentially abundant with DESeq2 and/or ANCOM in at least one of the two cohorts. Each box represents the first quartile, median and third quartile at the lower, middle and upper boundaries, with the whiskers representing points within 1.5 the interquartile range and the X representing the mean. Sample sizes: 131 PD and 84 HC subjects for the NL cohort and 55 PD and 87 HC subjects for the FIN cohort. NL cohort, Dutch case-control cohort; FIN cohort, Finnish case-control cohort; PD, Parkinson’s disease; HC, healthy control; DESeq2, Differential Expression analysis for Sequence count data; ANCOM, ANalysis of Composition of Microbiomes.

### DNA extraction methods

Samples were randomized as much as possible during the DNA extraction and library preparation and were all sequenced together to minimize batch effects (see Methods). Nonetheless, as the two different cohorts were initiated independently, different methods of sample collection and DNA extraction were used in the two cohorts. Samples from the NL cohort were collected without preservative, frozen immediately and processed using the Qiagen Allprep DNA extraction kit. In the FIN cohort, samples were collected using a DNA preservative and shipped via postal service. DNA extraction was performed using the PSP Spin Stool Kit. To assess the influence of the different methodologies, 47 stool samples from the NL cohort were collected and processed using both the NL method (Qiagen Allprep) and FIN method (PSP Spin Stool Kit) in parallel, taking one aliquot with each method from the same fecal material. Multivariate analysis yielded a statistically significant difference in overall microbiome composition between the two DNA extraction methods (PERMANOVA, p=9.999E-05). Several taxa at various taxonomic levels were identified as differentially abundant. Both ANCOM and DESeq2 identified various taxa belonging to the Lachnospiraceae family as less abundant when using the FIN extraction protocol, whereas DESeq2 also identified several taxa belonging to the order of Bacteroidales as more abundant when using the FIN protocol (Supplementary Table 6).

### Same household control subjects

Contrary to the FIN cohort, the NL cohort consisted of a substantial proportion of HC from the same household as a PD participant (n = 46). Similarity between same household participants was assessed by stratifying the NL cohort according to household status. Though the overall microbiome composition of age- and sex-matched non-household controls was more distant from PD subjects than that of non-matched same household controls, this did not lead to the discovery of differentially abundant taxa (Supplementary Table 7).

## Discussion

Here, we have presented the results of the two largest gut microbiome studies to date with treatment-naïve *de novo* PD subjects, close to the time of diagnosis. Both cohorts showed alterations in gut microbiome composition of fecal samples in PD after adjustment for relevant covariates. In concordance, several taxa at the ASV, genus and family level were identified as differentially abundant between PD and HC. Though none of the differentially abundant taxa could be directly replicated in both cohorts, both cohorts showed several ASVs and genera belonging to the family of Lachnospiraceae to be reduced in PD, in particular strains of the genus *Roseburia*.

### Comparison with previous studies

Previous gut microbiome studies in PD have consistently found differences in overall gut microbiome composition between PD and HC.^11–14^ The current study confirms the presence of gut microbiome alterations in treatment-naïve *de novo* PD. The contradictory findings on within-sample diversity (alpha diversity) can also be considered in line with the existing body of literature, with different studies finding either an increased or decreased alpha diversity in PD.^11^ Recently, a meta-analysis of previously published PD microbiome data concluded that differences in alpha diversity are not a marker of PD.^25^

Decreased levels of Lachnospiraceae and several of its taxa have been replicated in several PD microbiome studies.^11,14,15,19^, but have also been correlated to PD medication and more advanced disease stage.^14^ In our study several ASVs belonging to the genus *Roseburia*, as well as other genera belonging to the family Lachnospiraceae, were already decreased at the time of diagnosis before treatment initiation. This is in line with previous findings of reduced levels of Lachnospiraceae and two of its genera, including *Roseburia*, in a subcohort of 39 untreated *de novo* PD subjects, suggesting reduced levels of Lachnospiraceae are at least in part independent of PD medication and more advanced disease stage.^19^ Lachnospiraceae produce short chain fatty acids (SCFAs), known for their anti-inflammatory properties.^26^ Several studies have linked lower levels of SCFA-producing taxa with gut inflammation, epigenetic changes, and depressive symptoms in PD.^27–30^ In concordance, decreased levels of the SCFA-producing genera *Butyricoccus, Colidextribacter* and the families Butyricoccaceae and Veillonellaceae were found in the current study. On the other hand, the SCFA-producing family Rikenellaceae was increased in PD.

Levels of the non-SCFA-producing families Christensenellaceae and Eggerthellaceae were increased in one of the two cohorts. Increased levels of Christensenellaceae have been found in previous PD microbiome studies and seem in particular related to increased gut transit times.^31^ To our knowledge, Eggerthellaceae has not been identified as differentially abundant in previous human PD microbiome studies. Increased levels of *Bifidobacterium* and *Lactobacillus* were robustly replicated in several previous PD microbiome studies, but were hypothesized to be related to dopaminergic medication use.^14,18^ In accordance, no increased levels of *Bifidobacterium* were found in either cohort (NL cohort: reduced, nominal p=0.07; FIN cohort: increased, nominal p=0.14, Supplementary Table 4) nor of *Lactobacillus* (NL cohort: reduced, nominal p=0.92; FIN cohort: increased, nominal p=0.99, Supplementary Table 4). Increased levels of the genus *Akkermansia* is one of the most replicated taxonomic differences between PD and HC.^11,15^ In addition, *Akkermansia* is increased in RBD-positive probable prodromal PD subjects.^22^ *Akkermansia* is hypothesized to aggravate gut inflammation, increase gut permeability and is associated with levels of constipation.^32^ In particular, *Akkermansia muciniphila* is known to thrive in fiber-depleted environments as it uses mucin as energy source, thereby degrading the intestinal wall and leaving the host more vulnerable to epithelial access of intestinal pathogens.^33^ However, the current study found no evidence of increased levels of *Akkermansia* (Supplementary Table 4).

Fewer taxonomic differences were found in the current cohorts than reported in most previous PD microbiome studies.^11^ This stresses the need for treatment-naïve *de novo* cohorts to avoid the putative confounding effects of disease duration, deteriorating health and dopaminergic treatment. Adjusting for relevant confounders in the current study might have further filtered out non-PD specific taxa. In particular, constipation is a well-known non-motor symptom of PD and important determinant of gut microbiome composition.^34,35^ Previous studies used questionnaires or single questions to assess constipation, but these methods are notorious for their low ability to detect constipation in PD.^35,36^ Here, stool diaries measuring stool frequency and stool consistency were used, possibly providing a more objective and adequate adjustment for constipation.^37,38^

### Differences between cohorts

Though both cohorts showed several differentially abundant taxa belonging to the family of Lachnospiraceae, and the direction of the difference for differentially abundant taxa was often the same, it is striking that no taxon could be replicated in both cohorts with statistical significance. Though previous studies also reported various inconsistent results, certain study characteristics might explain inconsistencies between the two cohorts. First, the NL control cohort consisted of a large number of spouses from participants. Microbiome composition was less distant between spousal cases and controls. In addition, non-spousal controls were often spouses of PD patients from the Groningen PD expertise center who did not participate in the study. Possibly, a shared living environment with PD subjects might have ameliorated differences in gut microbiome composition in the entire NL cohort. Second, the different DNA extraction methods affected the differentially abundant taxa, in particular of taxa belonging to the family of Lachnospiraceae. Third, the recruitment differed between the two cohorts. Patients were directly referred by a neurologist for the purpose of participating in a PD study in case of the NL cohort. In the FIN cohort patients were selected after referral for a DAT-SPECT scan due to diagnostic uncertainty, possibly leading to an overrepresentation of untypical phenotypes. Fourth, the genetic background of the two cohorts might differ, with the Finnish population being particularly isolated in Europe, possibly driving different host-microbiome interactions.^18,39,40^ Last, though no dietary variable was selected as a significant confounder for each cohort separately, no direct comparison of diet was possible between the two cohorts. Therefore, the NL and FIN cohorts might represent different dietary habits.

## Limitations

The current study has some clear advantages over previous microbiome studies in PD, such as the inclusion of treatment-naïve *de novo* PD subjects, the use of two separate cohorts and a more adequate assessment of diet and constipation. Nonetheless, a few limitations need to be addressed. First, two different DNA extraction methods were used in both cohorts. As discussed, differences in gut microbiome composition driven by the DNA extraction method were assessed by parallel sampling of 47 stool samples using both methods. However, DNA extraction should ideally be performed using the same method for a direct comparison between cohorts.

Second, samples were randomized as much as possible during the DNA extraction and library preparation and were all sequenced together to minimize batch effects. Since already extracted HC samples from a previous publication were used to supplement the HC group of the FIN cohort, these samples could not be randomized during DNA extraction. In the NL cohort the DNA extraction batch was marked as a relevant confounder, but did not have a significant association in the final multivariable model (Table 2). In addition, batch effects from DNA extraction are less likely to impact the results of high-biomass samples such as feces.^41^ However, some confounding in the FIN cohort cannot be excluded.

Third, despite presenting the two largest gut microbiome cohorts in *de novo* PD thus far, failed replication might still result from a lack of power. Provided a confounding effect of dopaminergic medication and the association of longer disease duration with more pronounced changes in gut microbiome composition, smaller effect sizes are expected in a treatment-naïve *de novo* PD cohort. Arguably, at least at the taxonomic levels of genus and higher, the gut microbiome seems unsuitable as a diagnostic biomarker, although this should ideally be compared to relevant differential diagnoses or re-evaluated in relation to potential PD subtypes. Nonetheless, a lack of power might still obscure pathophysiological relevant associations. Wallen et al. were the first to describe overabundance of the putative opportunistic pathogens *Porphyromonas, Prevotella* and *Corynebacterium* at the genus level and attributed these novel findings to the larger sample size compared to previous studies.^14^ Overabundance of these previously reported opportunistic taxa was not found in the current cohorts, but might be missed due to insufficient power.

Last, participants were excluded if they had used antibiotics in the month before stool sample collection. Even though antibiotics induced gut microbiota changes most often subside within 28 days, changes might persist for as long as several years, depending on the antibiotic used and inter-personal variability.^42^ A small bias can therefore not be excluded.

### Future perspective

Expansion of the current dataset can further confirm the association between PD medication and gut microbiota. Though the current study cannot be confounded by the effect of PD medication, no causal inferences can be made based on the absence of previously described gut microbiome changes associated with PD medication use. Levodopa is suggested to drive positive selection of tyrosine decarboxylase producing bacteria, whereas overall dopaminergic input might influence gut microbiome composition via cerebral signaling or modulating stool transit times.^43,44^ Currently, only one study investigated the influence of Levodopa initiation on gut microbiome composition in PD, but found no associated changes.^45^ This study, however, only included 19 participants and included several participants who were stable users of other dopaminergic medication. Follow-up should ideally be done within two years, as gut microbiome composition does not seem to shift significantly as a result of disease progression in a two-year period.^13^ This might confirm the association between dopaminergic medication and *Bifidobacterium, Lactobacillus* and tyrosine decarboxylase producing bacteria.

Extended follow-up after two years would make it possible to analyze the association between disease progression and increased gut microbiome composition changes, particularly further decrease of SCFA-producing taxa and possible increases in opportunistic pathogens. Additionally, adequate subtyping could lead to the identification of subtype-specific microbiome changes. Given the large clinical heterogeneity of PD, different subtypes might represent different pathophysiological mechanisms and ports d’entrée. The recently proposed dichotomy of brain-first versus body-first PD might be of particular interest, with gut microbiome changes possibly being more pronounced in a body-first subtype.^7^ The body-first subtype is currently defined by polysomnography proven RBD at the time of diagnosis and/or during the prodromal stages of PD. Questionnaire based assessments of RBD have a far lower sensitivity and specificity in *de novo* PD, and a lower predictive value in case of probable prodromal PD.^23,46^ Therefore, adequate biomarkers of the body-first subtype, besides polysomnography, remain to be established.

Though taxonomic research can already provide insight into the relationship between PD and gut microbiota, integration with data on the functionality of the intestine and the gut microbiome, could further elucidate relevant mechanisms. These include shotgun metagenomics and metabolomics studies, as well as markers of gut permeability, intestinal inflammation and systemic inflammation. Shotgun metagenomic sequencing would provide complete taxonomy data at the species level and would additionally provide data on the metabolic pathways associated with the sequenced gene fragments.^48,49^ Metabolomic analysis would provide actual functional readouts of the gut microbiome. Current data on the metabolome of gut microbiota suggests reduced levels of SCFAs, which would be in line with previous taxonomic studies and our current findings.^28^ However, metabolomic data of treatment-naïve *de novo* PD subjects is still lacking and could generate valuable hypotheses to be assessed in pre-clinical functional studies. Data on gut permeability, intestinal inflammation and systemic inflammation can further indicate the means through which gut microbiota might increase vulnerability to PD pathology.^47^

## Conclusion

Our findings suggest fewer taxonomic differences in treatment-naïve *de novo* PD subjects compared to previous studies with already treated patients in the more advanced stages of the disease. Although our finding of reduced levels of SCFA-producing taxa are in line with previous studies, none of our findings could be directly replicated with statistical significance in both cohorts, showing the importance of comparing multiple cohorts for analyzing highly variable data such as the microbiome. In addition, the importance of adequately assessing the effects of potential nuisance variables, such as constipation and batch effects of laboratory procedures, can be highlighted. Further enquiry of the metabolic profile of the gut microbiome in treatment-naïve *de novo* PD and longitudinal analysis of the current dataset can provide further insight in relevant pathophysiological mechanisms and the extent to which gut microbiome changes might be confined to specific PD subtypes and medication effects.

## Methods

### Study population

For the Dutch (NL) cohort, treatment-naïve *de novo* PD participants were included as part of the Dutch Parkinson Cohort of de novo PD subjects (DUPARC, ClinicalTrials.gov identifier NCT04180865).^48^ Inclusion criteria were PD diagnosis by a movement disorder specialist according to the Movement Disorders Society (MDS) clinical diagnostic criteria,^1^ confirmed by a dopaminergic deficit quantified by FDOPA-PET or one-year follow-up if no FDOPA-PET was performed (n=8). HC did not have a neurodegenerative disorder and could not be classified as probable prodromal PD.^4^ HC were recruited from the same geographical area and were spouses of PD participants, caregivers of PD patients at the PD expertise center Groningen, or respondents to local advertisements.

For the Finnish (FIN) cohort, treatment-naïve *de novo* PD participants were included as part of the Non-Motor Symptoms and DopAmine Transporter binding study (NMDAT study, ClinicalTrials.gov identifier NCT02650843). The patients were scanned with [I-123]FP-CIT SPECT because of parkinsonism or tremor for which they were referred to imaging by their neurologist. Inclusion criteria for this microbiome study were PD diagnosis by a movement disorder specialist according to the MDS clinical diagnostic criteria,^1^ confirmed by a dopaminergic deficit quantified by [I-123]FP-CIT SPECT. [I-123]FP-CIT SPECT were analyzed with BRASS software (Hermes Medical Solutions AB, Stockholm, Sweden), in which a dopaminergic deficit was defined as more than two standard deviations below the reference mean in any of the six analyzed regions. The study subjects were required to be aged 18 or over and to be able to understand and answer the questionnaires in Finnish. Only the patients who also filled out the gastrointestinal questionnaires were included in the current analyses. HC samples of the FIN cohort were from participants of the previously described PD microbiome follow-up study by Aho et al.^13^ and HC of the GAMbling and DopAmine Transporter binding (GAMDAT) study.^49^

Shared exclusion criteria for both cohorts comprised major confounders of gut microbiome composition (eg. recent antibiotics usage in the previous month). A complete overview of the exclusion criteria per study is provided in the Supplementary Methods. All studies were approved by their respective local ethics committee and all participants provided written informed consent.

### Data and stool sample collection

Participants in the NL cohort collected their stool sample at home and stored it in their home freezer (−20°C). Stool samples were then stored at -80°C at the University Medical Center Groningen (UMCG) and were kept frozen on dry ice during all transportation. PD participants in the NL cohort were extensively characterized, as previously described,^48^ with HC receiving a more selective assessment. For the current study, determinants of gut microbiome composition were selected as potential confounders in three different categories: (1) the medical history, including disease history, medication use (eg. use of proton pump inhibitors, opioids, antidepressants) and family history; (2) nutrient intake based on a dietary diary that was filled out for three consecutive days, including alcohol usage; (3) gastrointestinal functioning represented by stool consistency and frequency based on a stool diary that was filled out for seven consecutive days, with consistency represented by the Bristol Stool Chart, and the subjective sense of constipation asked with question 5 of the Non-Motor Symptom Questionnaire (NMSQ).^50^ In addition, motor-symptomatology was examined using the Movement Disorder Society Unified Parkinson Disease Rating Scale (MDS-UPDRS) part III.^51^

Participants in the FIN cohort collected their stool sample at home using a collection tube with DNA-stabilizing buffer (PSP Spin Stool DNA Plus Kit, STRATEC Molecular), which were send via postal services to the University of Helsinki laboratory where they were stored at -80°C. If samples were not immediately shipped, they were stored at home in the refrigerator. Participants in the FIN cohort received similar assessments of potential confounders of gut microbiome composition and PD status in the categories of medical history, diet and gastrointestinal function. Notable differences compared to the NL cohort were the usage of a food frequency questionnaire, concerning food consumption in the month before stool sampling, instead of a dietary diary. Also, there were several additional measures of subjective constipation instead of question 5 of the NMSQ: a question about strained defecation in the stool diary and the ROMEIII, Wexner and Constipation Severity Index (CSI) questionnaires. A complete overview of the assessed endpoints per cohort is provided below in Supplementary Methods Table 1.

### Laboratory procedures

All laboratory procedures were performed at the Institute of Biotechnology, University of Helsinki. DNA extraction of the NL samples was performed using the Qiagen Allprep DNA/RNA Mini Kit, whereas the PSP Spin Stool DNA Plus Kit (Invitek Molecular, Germany) was used for the FIN samples. To assess the possible influence of both sample collection and DNA extraction protocols, a subset of the NL participants (n=47) collected an additional stool sample from the same feces as their regular stool sample collection. The additional sample was collected using the FIN cohort protocol with PSP tubes and processed using the PSP Spin Stool Kit. The V4 hypervariable region of the 16S rRNA gene was amplified using a mixture of universal primers 515F1-4 (5’-GTGCCAGCMGCCGCGGTAA-3’) and 806R1-4 (5’-GGACTACHVGGGTWTCTAAT-3’) with partial Illumina TruSeq adapter sequences added to the 5′ ends: F1 ACACTCTTTCCCTACACGACGCTCTTCCGATCT, F2 ACACTCTTTCCCTACACGACGCTCTTCCGATCTca, F3 ACACTCTTTCCCTACACGACGCTCTTCCGATCTgca, F4 ACACTCTTTCCCTACACGACGCTCTTCCGATCTagcaatt, R1 GTGACTGGAGTTCAGACGTGTGCTCTTCCGATCT, R2 GTGACTGGAGTTCAGACGTGTGCTCTTCCGATCTca, R3 GTGACTGGAGTTCAGACGTGTGCTCTTCCGATCTatct, R4 GTGACTGGAGTTCAGACGTGTGCTCTTCCGATCTtctact

The additional nucleotides (non-capitalized letters) are introduced for mixing in sequencing. The 2-step PCR amplification was done as described in Aho et al 2019.^13^ Every DNA extraction and amplification batch included a blank control to assess possible contamination and case and control samples were semi-randomized, where possible, to avoid batch effects. Barcodes were selected using BARCOSEL.^52^ All samples were sequenced together for four runs using Illumina MiSeq (v3 600 cycle kit), with 325 bases for the forward and 285 bases for the reverse read.

### Bioinformatics

Forward and reverse primers were removed from R1 and R2 reads using cutadapt v2.10.^53^ Further bioinformatics were performed with DADA2 v1.18.^54^ Forward sequences were trimmed at 200 nucleotides (nt) and reverse sequences at 150nt, quality trimmed at the first instance of a nucleotide (or group of nucleotides) with quality score 2. Reads with more than 10 expected errors or ambiguous nucleotides were discarded. Amplicon sequence variants (ASVs) were inferred for each of the four runs separately, using default parameters in DADA2. An overview of the number of sequences per sample during each of the preprocessing steps (primer removal, quality filtering and length trimming, denoising forward and reverse reads, merging forward and reverse reads) is provided in Supplementary Table 8. In addition, the number of sequences of the blank control samples at the DNA extraction and PCR steps are included, showing very minimal to no contamination during the wet lab work (Supplementary Table 8). Subsequently, the four resulting sequence tables were merged, ASVs that only differed in length were collapsed and chimera were removed. Taxonomic assignments were based on the SILVA v138 reference database,^55^ in which taxonomic assignments are first performed up to genus level using a naïve Bayesian classifier with 50% bootstrap confidence. Second, species are assigned based on a 100% match with the references sequence if the match corresponds to the already assigned genus.

### Statistical analysis

Due to the geographical and cultural differences, as well as the different endpoints that were assessed, the NL and FIN cohort were analyzed separately. Supporting the large influence of geographical differences, samples were more distant in terms of their overall microbiome composition when contrasting the geographical origin, compared to PD and HC status (Supplementary Methods Figure 2). All statistical analyses were performed with the statistical software R, v4.0.3. The threshold for statistical significance was set at p < 0.05, or adjusted p < 0.05 if corrected for multiple comparisons. Comparison of clinical and technical variables were performed using a Chi-square test for categorical variables and a Student’s t-test or a Mann-Whitney U test for continuous variables, depending on the distribution of the variable. For microbiota analyses, the sequence tables, taxonomy tables and metadata tables were merged in a phyloseq object (phyloseq, v1.34.0). Multivariate analyses of overall community structure were performed on Aitchinson distances: Euclidean distances calculated after centered log-ratio (clr) transformation of the ASV count data.^56^ Statistical significance was calculated using a PERMANOVA with adjustment for selected variables (see variable selection).^57^ PERMANOVA was performed with marginal testing, meaning all variables were assessed whilst holding constant all other variables in the model. In parallel, a non-compositional approach was performed using Bray-Curtis distances after rarefaction.^56^ Cut-offs for rarefaction were identified for both datasets separately, based on the plateauing of the rarefaction curves depicting sample richness (Supplementary Methods Figure 1). Similarly, the differential abundance analyses were performed with adjustment for selected confounders using both a compositional approach with ANCOM v2.1 and a non-compositional approach with DESeq2.^58–60^ Differential abundance analyses were performed at the ASV, genus and family level after filtering out taxa which occurred in fewer than ten percent of the samples. Taxa were identified as differentially abundant with a detection threshold of 0.7 in ANCOM or with a false discovery rate (FDR) adjusted p-value < 0.05 in DESeq. Alpha diversity was investigated using alpha diversity indices Chao1, Shannon, Inverse Simpson and observed richness, which were tested univariably using a Mann-Whitney U test.

### Variable selection

Given the primary aim to describe gut microbiome changes for the first time in treatment-naïve de novo PD subjects, all clinical data described under “data and stool sample collection” and technical metadata (eg. DNA extraction batch, PCR batch, number of reads) were investigated for their potential to influence the effect of group status on gut microbiome composition. The variables age, sex, BMI, stool consistency and DNA extraction batch were selected a priori as potentially relevant nuisance variables. To ensure a large effect of another variable on the relation between PD status and gut microbiome composition would not be missed, all variables were screened for their potential effect using a PERMANOVA in a model with PD status:

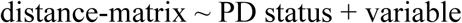

Variables that shifted the explained variance of PD status (R2) by at least ten percent compared to the univariable explained variance of PD status (distance-matrix ∼ PD status), were added to the list of relevant model covariates (Supplementary Table 1). Subsequently, the selection of relevant covariates was investigated for collinearity with PD status. Variables with a generalized variance inflation factor (GVIF) ≥2 with PD status were excluded. In the overall model, a GVIF of 3 was used as threshold for collinearity, excluding variables with a higher GVIF or one of several variables that only showed collinearity amongst each other (Supplementary Table 2). This resulted in the following models for the multivariate analysis of overall gut microbiome composition.

For the NL cohort: distance-matrix ∼ PD status + age + sex + BMI + stool consistency + NMSQ constipation + stool frequency + DNA extraction batch

For the FIN cohort: distance-matrix ∼ PD status + age + sex + BMI + stool consistency + CSI total score + strained defecation + number of sequences

For the differential abundance analysis, covariates were selected that showed a significant relationship with the overall gut microbiome composition in the final model with p < 0.1.

For the NL cohort: taxon ∼ PD status + BMI + stool consistency + stool frequency For the FIN cohort: taxon ∼ PD status + stool consistency + number of sequences

### Data availability

Sequencing data of the V4 region of the 16S rRNA gene will be made available via the European Nucleotide Archive upon publication. Sample metadata can be requested from the respective principal investigator of DUPARC, NMDAT, GAMDAT or the Aho et al. 2019 baseline and follow-up dataset via the corresponding author upon reasonable request, depending on associated privacy regulations.

## Funding sources

DUPARC: Selfridges Group Foundation (Weston Brain institute).

NMDAT: Turku University Hospital (VTR-funds), Päivikki and Sakari Sohlberg Foundation, Finnish Cultural Foundation (Varsinais-Suomi regional fund), Turku University Foundation, The Academy of Finland (295724, 310835), Hospital District of Helsinki and Uusimaa (UAK1014004, UAK1014005, TYH2018224), Finnish Medical Foundation, Finnish Parkinson Foundation, Maire Taponen Foundation.

GAMDAT: The Finnish Foundation for Alcohol Studies.

We thank all participants in the NL and FIN cohort. For their aid in the recruitment of participants for the NL cohort, we thank the Parkinson Platform Northern Netherlands (PPNN) Study Group collaborators.* We would like to thank Renée Speijers, Yvonne Nijman and Hanna Slomp for their help in the recruitment and logistics of the study. For the FIN cohort we thank drs. Antti Loimaala and Sorjo Mätzke as well as the teams of the nuclear medicine departments in Meilahti and Jorvi hospitals for allowing us to recruit and study patients in their departments and supporting us throughout the study. We thank Kari Lindholm for the help in the data collection. We thank Velma Aho for her work on the Finnish gut microbiome PD follow-up study and her advice concerning data analysis.

We acknowledge the DNA Sequencing and Genomics Laboratory, Institute of Biotechnology, University of Helsinki, for DNA isolations and sequencing of FIN and NL cohort samples, in particular Eevakaisa Vesanen, Ursula Lönnqvist and Julia Teleni.

*PPNN Study Group

Verwey NA^1^, Van Harten B^1^, Portman AT^2^, Langedijk MJH^2^, Oomes PG^2^, Jansen BJAM^2^, Van Wieren T^2^, Van den Bogaard SJA^3^, Van Steenbergen W^3^, Duyff R^3^, Van Amerongen JP^3^, Fransen PSS^4^, Polman SKL^4^, Zwartbol RT^4^, Van Kesteren ME^4^, Braakhekke JP^4^, Trip J^4^, Koops L^4^, De Langen CJ^4^, De Jong G^4^, Hartono JES^4^, Ybema H^4^, Bartels AL^5^, Reesink FE^5^, Postma AG^6^, Vonk GJH^7^, Oen JMTH^7^, Brinkman MJ^7^, Mondria T^7^, Holscher RS^7^, Van der Meulen AAE^8^, Rutgers AWF^8^, Boekestein WA^9^, Teune LK^9^, Orsel PJL^10^, Hoogendijk JE^10^, Van Laar T^11^.

^1^Department of Neurology, Medisch Centrum Leeuwarden, Leeuwarden, the Netherlands; ^2^Department of Neurology, Treant Zorggroep locations, Stadskanaal, Emmen, Hoogeveen, the Netherlands; ^3^Department of Neurology, Tjongerschans Ziekenhuis Heerenveen, Heerenveen, the Netherlands; ^4^Department of Neurology, Isala, Zwolle, Meppel, the Netherlands; ^5^Department of Neurology, Ommelander Ziekenhuis Groningen, Scheemda, the Netherlands; ^6^Department of Neurology, Nij Smellinghe Ziekenhuis Drachten, Drachten, the Netherlands; ^7^Department of Neurology, Antonius Zorggroep, Sneek, the Netherlands; ^8^Department of Neurology, Martini Ziekenhuis, Groningen, the Netherlands; ^9^Department of Neurology, Wilhelmina Ziekenhuis Assen, Assen, the Netherlands; ^10^Department of Neurology, Sionsberg, Dokkum, the Netherlands; ^11^Department of Neurology, University Medical Center Groningen, Groningen, the Netherlands.

## Authors contributions

JMB contributed to the study conception, participant inclusion, data collection, data analysis, writing the first draft of the manuscript and later reviewing of the manuscript. KM contributed to study conception, participant inclusion, data collection, writing the first draft of the manuscript and later reviewing of the manuscript. PABP contributed to data analysis and reviewing of the manuscript. SvdZ contributed to participant inclusion, data collection and reviewing of the manuscript. THM contributed to data collection and reviewing of the manuscript. RL contributed to data collection and reviewing of the manuscript. TN contributed to data collection and reviewing of the manuscript. PL contributed to data analysis and reviewing of the manuscript. LP contributed to the design of the laboratory work, data analysis and reviewing of the manuscript. EM contributed to data collection and reviewing of the manuscript. EJ contributed to data collection and reviewing of the manuscript. EP contributed to data collection and reviewing of the manuscript. VK contributed to participant inclusion, data collection and reviewing of the manuscript. PA contributed to the design of the laboratory procedures, data analysis and reviewing of the manuscript. FS contributed to the study conception, participant inclusion, data collection and reviewing of the manuscript. TvL contributed to the study conception, participant inclusion, data collection and reviewing of the manuscript.

