## Supplementary Methods for "Gut microbiome alterations in fecal samples of treatment-naïve *de novo* Parkinson’s disease patients"

**Contents**

1. Exclusion criteria
2. Supplementary methods table 1: overview of assessed endpoints
3. Supplementary methods figure 1: rarefaction curves
4. Supplementary methods figure 2: Overall microbiome composition of both cohorts according to (A) group (PD and HC) status and (B) country of origin.

**Exclusion criteria NL cohort**

All participants:

- Active or persistent primary disease of the gastrointestinal tract
- History of peritonitis, severe endometriosis, abdominal, intestinal or urogenital fistula,
- Hepatobiliar or pancreatic disease (except asymptomatic cholecystolithiasis)
- History of abdominal or anorectal surgery, except minor surgery such as uncomplicated appendectomy or cholecystectomy (>6 months ago).
- Severe gynaecological prolapse (grade III)
- Cancer and/or adjuvant treatment within the last 6 months
- Within the last three months: severe hypo- or hyperkalemia, narcosis, analgosedation, endoscopic procedure of the gastrointestinal tract, abdominal trauma
- Within the last three months: gastrointestinal tract infection, food intoxication
- Antibiotic use in the last month

PD participants:

- History of dopaminergic medication use

HC participants:

- History of neurodegenerative disease, in particular signs of parkinsonism.
- Probable prodromal PD^5^

**Exclusion criteria FIN cohort**

All participants NMDAT:

- Any kind of limitation affecting their ability to understand the informed consent, such as significant mental health problems or cognitive problems (Mini-Mental State Examination (MMSE)<18

PD participants NMDAT:

- History of dopaminergic medication use
- Antibiotic use in the last month
- Imaging with a scanner whose binding ratios cannot be compared to other scanners´ values

HC participants GAMDAT:

- Evidence of clinically significant cardiovascular, renal, hepatic, hematological, gastrointestinal, pulmonary, endocrinological or neurological disease
- Evidence of current alcohol or substance use disorder within last 6 months (DSM-V)
- Intoxicated or recent (less than 36 h) drug or alcohol usage
- Body weight > 180 kg (scanner limit)
- Strong susceptibility to allergic reactions or nausea
- Blood donation within 60 days prior to the study
- Has undergone a prior PET or SPECT study
- Current treatment with amphetamine derivatives, methylphenidate, bupropion or other medications known to interfere with DAT imaging if the medication cannot be stopped for a minimum of 1 month before DAT imaging
- Any contraindication to magnetic resonance imaging (MRI)
- Current pregnancy (i.e. has to be surgically sterilized, at least one year postmenopausal, or negative serum pregnancy test before the study) or lactation in women
- Current psychiatric DSM-V Axis-I disorder (e.g. major depression, bipolar disorder psychoses)

HC participants Aho et al (assessed at baseline inclusion for Scheperjans 2015^6^):

- Active smoking in last 6 months
- Diagnosis of dementia or MMSE < 25 points
- Diagnosis of major depression or GDS-15 > 9 points
- Diagnosis of psychosis
- Any signs of parkinsonism
- Hyposmia
- REM sleep behaviour disorder
- Restless legs syndrome
- NMSQuest score >3 (not including items 4, 6, 7, 10, 18, 23, 27)
- First degree relative or more than one relative with PD
- HIV infection
- Living in same household with PD patient
- Active or persistent primary disease of gastrointestinal tract: e.g. celiac disease, pernicious anemia, autoimmune gastritis, symptomatic diverticulosis, inflammatory bowel disease, irritable bowel syndrome, strictures, adhesions, varicosis or diverticulum of the esophagus, Meckel’s diverticulum (Exception: other forms of chronic gastritis)
- Endocrinological disease (Exception: diabetes mellitus without polyneuropathy; treated hypothyreosis with normal thyrotropin level)
- Alcohol abuse
- B-hypovitaminosis
- History of hepatobiliar or pancreatic disease (Exception: asymptomatic cholecystolithiasis)
- Previous abdominal or anorectal surgery (Exceptions: can be enrolled
- after uncomplicated haemorrhoid procedure and 1 year after uncomplicated appendectomy, inguinal hernia repair, cholecystectomia, or gynecologial surgery if symptoms resolved and no signs of adhesions)
- Severe gynaecological prolapse (grade III) (Exception: can be enrolled after repair procedure if symptoms resolved and no signs of adhesions)
- History of peritonitis, severe endometriosis, polyneuropathy, polio, spina bifida, severe symptomatic spinal stenosis (if not symptom free for at least 1 year), paraparesis of any cause, symptomatic peripheral arteriosclerosis, any signs or history of intestinal ischemia (e.g. claudication), aortic aneurysm or dissection, connective tissue disease, autoimmune disease, sarcoidosis outside of the lungs or skin, cancer that is not considered cured, abdominal, intestinal, or urogenital fistula (Exceptions: treated autoimmunehypothyreosis with normal thyrotropin level)
- Heart failure (Exception: isolated left ventricular heart failure NYHA ≤ II)
- Known severe renal insufficiency (glomerular filtration rate < 30 ml/min)
- One of the following within the previous 2 months: severe hypokalemia or hyperkalemia demanding hospital treatment, narcosis or analgosedation, endoscopic procedure of the gastrointestinal tract, abdominal trauma
- Any of the following within the last 2 months: gastrointestinal or respiratory tract infection, food intoxication, major epistaxis requiring treatment by a physician
- Antibiotic treatment within the last month
- Any drug abuse
- Any regular use (>2 times a week) of the following medications over the last 2 months: opioids, loperamide, inhaled β-agonists or anticholinergics, glucocorticoids (oral or parenteral), tricyclic antidepressants, antihistaminics with systemic anticholinergic effects, metoclopramide, cholinergics, anticholinergics (except for PD indication), domperidone, protone pump inhibitors

| **Supplementary methods table 1:** overview of assessed endpoints | | | | | |
| --- | --- | --- | --- | --- | --- |
| **Variable** | **NL cohort PD (DUPARC)** | **NL cohort HC (DUPARC)** | **FIN cohort PD (NMDAT)** | **FIN cohort HC (GAMDAT)** | **FIN cohort HC (Aho et al.)** |
| **Technical variables** |  |  |  |  |  |
| DNA extraction method | X | X | X | X | X |
| DNA extraction batch | X | X | X | X | X |
| PCR batch | X | X | X | X | X |
| Number of reads | X | X | X | X | X |
| **Medical history** |  |  |  |  |  |
| Age | X | X | X | X | X |
| Sex | X | X | X | X | X |
| BMI | X | X | X | X | X |
| Disease history | X | X | X | X | X |
| Medication use | X | X | X | X | X |
| Family anamnesis | X | X | X | X | X |
| Allergies | X | X | X | X | X |
| Lactose intolerance | X | X | X | X | X |
| Alcohol use | X | X | X | X | X |
| Smoking | X | X | X | X | X |
| Drug use | X | X | X | X | X |
| **Gastrointestinal health** |  |  |  |  |  |
| Stool diary | X | X | X | X | X |
| Bristol Stool Chart | X | X | X* | X* | X* |
| RomeIII |  |  | X | X | X |
| Constipation Severity Index |  |  | X | X | X |
| Wexner |  |  | X | X | X |
| NMSQ question 5: constipation | X | X |  |  |  |
| **Dietary habits** |  |  |  |  |  |
| Dietary diary | X | X |  |  |  |
| Food frequency questionnaire |  |  | X | X | X |
| **PD clinical characteristics** |  |  |  |  |  |
| MDS-UPDRS part III | X | X | X |  |  |
| Hoehn and Yahr | X | X | X |  |  |
| MDS-UPDRS, Movement Disorder Society Unified Parkinson Disease Rating Scale; NMSQ, non-motor symptom questionnaire.  *A modified version of the Bristol Stool Chart was used in the FIN cohort | | | | | |

**Supplementary methods figure 1:** rarefaction curves

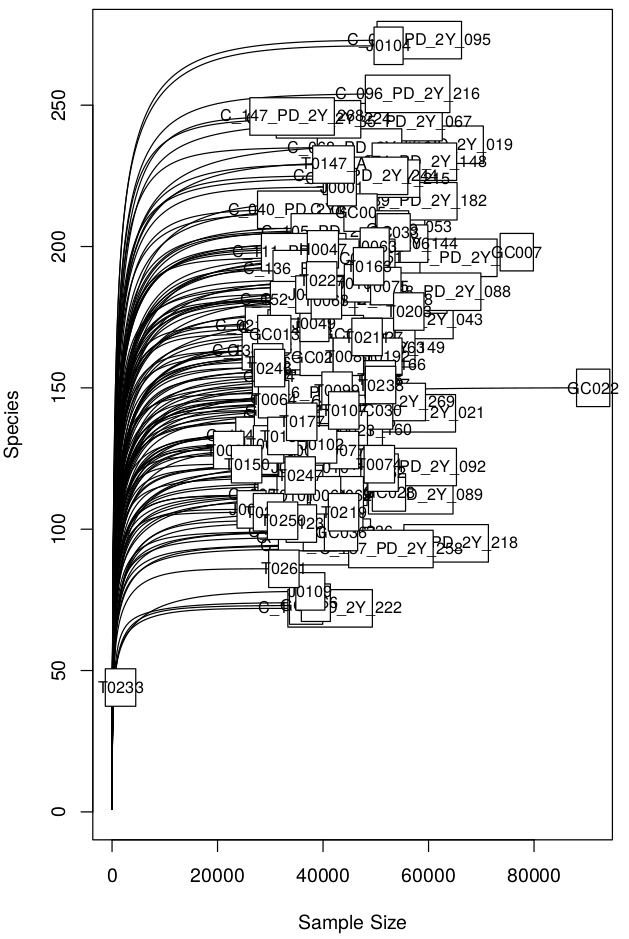

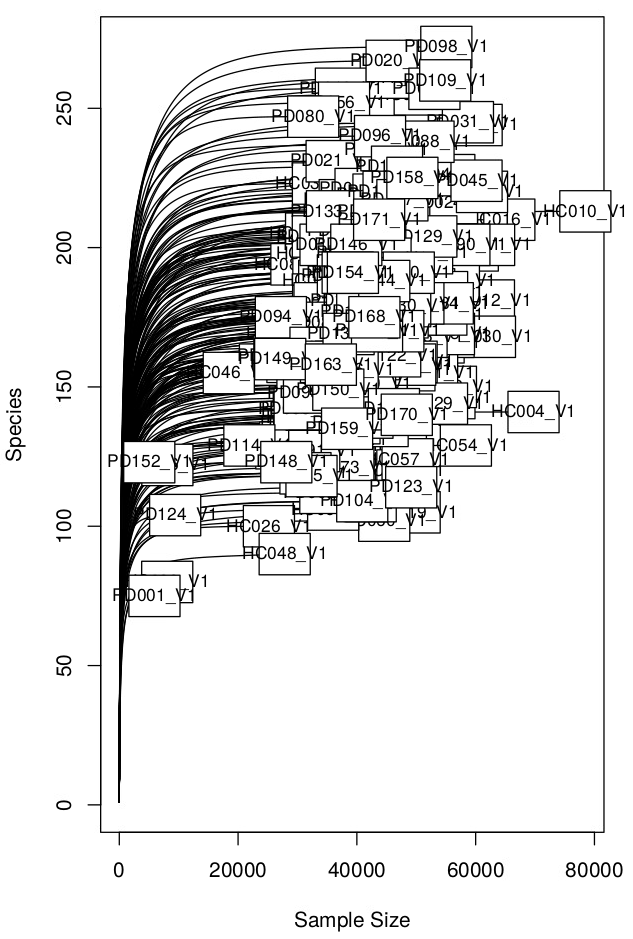

**Supplementary methods figure 2**

A.
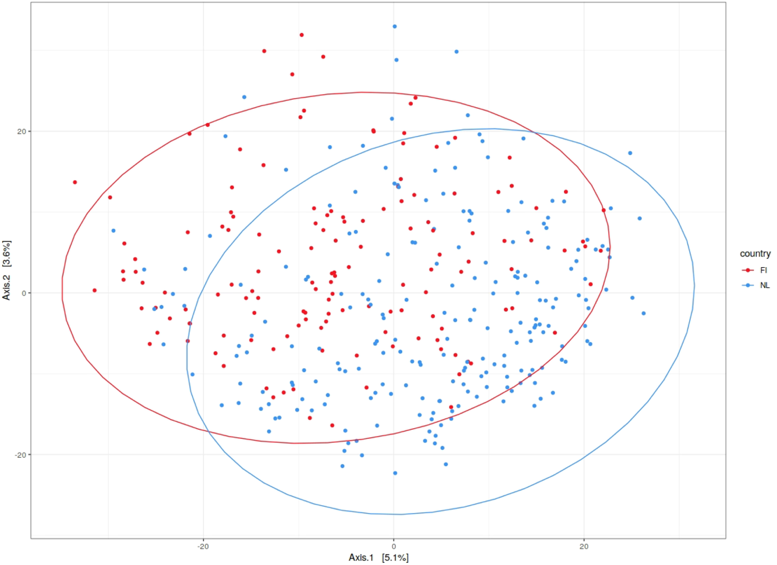
**Group	B. Country**

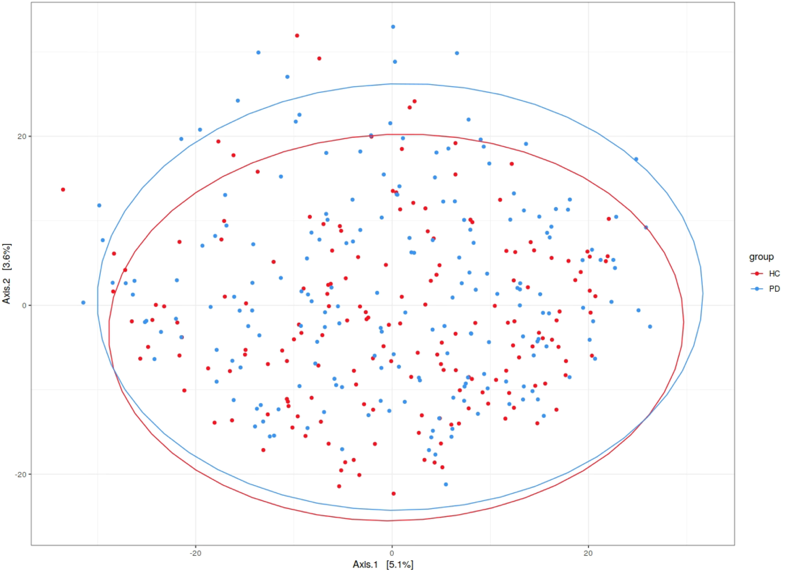

Overall microbiome composition of both cohorts according to (A) group (PD and HC) status and (B) country of origin. A larger distance between the overall microbiome composition of the two different countries can be observed compared to group status. This was also confirmed by means of PERMANOVA using 1E6 permutations: group status R2 = 0.00534, p = 9E-06; country R2 = 0.02218, p = 1E-06 (lowest possible p-value with 1E6 permutations).
